# Health-related quality of life in mild-moderate patchy alopecia areata: Results from the first controlled Phase 2 clinical trial in this population with STS01 (dithranol/ProSilic) and challenges for the future

**DOI:** 10.64898/2026.04.02.26349940

**Authors:** Kerry Montgomery, Andrew Messenger, David Fleet

## Abstract

A phase 2, randomised, double-blind, placebo-controlled trial has been conducted in mild-to-moderate patchy alopecia areata (AA). This demonstrated significant and dose-related improvements in hair regrowth with STS01, a controlled release, topical formulation of dithranol. Here we report the results of the Alopecia Areata Symptom Impact Scale (AASIS) that assesses the severity of symptoms, daily functioning and feelings. Similar to trials in severe AA, significant improvements in hair regrowth did not translate into significant health-related quality of life (HRQoL) improvements, even in patients with complete hair regrowth, although there was some treatment-related correlation between changes in AASIS scores from baseline and clinical assessment SALT scores. The use of current HRQoL methods or indeed new measures in development for future trials, will have considerable challenges: patients may not have a ‘true’ baseline at entry, may develop coping mechanisms, and there may be a delay between physical and psychological improvement.

**Learning points:**

- Alopecia areata is an autoimmune disease that imposes a substantial psychosocial burden on individuals.
- Poor health-related quality of life (HRQoL) is not directly related to the severity of hair loss as measured by SALT score. Patients with mild to moderate AA (≤50% hair loss as measured by SALT score) are similarly affected to those with severe AA (SALT score >50)
- STS01 is a novel controlled release, topical formulation of dithranol that showed significant and dose-related improvements in hair regrowth compared to placebo in a phase 2 double-blind randomised clinical trial in patients with mild to moderate AA.
- However, similar to trials in severe AA, significant improvements in hair regrowth did not appear to translate into significant HRQoL improvements, as measured using the Alopecia Areata Quality of Life Questionnaire.
- There are substantial limitations in the current tools for measuring HRQoL in individuals with AA, highlighting the need for increased understanding of the impact and determinants of HRQoL to help address these challenges.

## Background

Alopecia areata (AA) is a chronic autoimmune condition affecting the hair follicles, characterized by sudden and unpredictable hair loss. Hair loss ranges from patchy scalp hair loss to loss of all scalp and body hair. JAK inhibitors have recently been licenced for severe AA, but there are no treatments licenced for patients with mild to moderate AA (encompassing <50% of the scalp). AA imposes a substantial psychosocial burden on individuals,^1^ and treatment goals should include improving health-related quality of life (HRQoL). HRQoL is impaired in patients with mild to moderate AA to a similar extent as with severe AA.^2^

STS01 is a controlled-release, topical formulation of dithranol incorporating Prosilic, a novel silicon-based nanoparticle controlled-release mechanism. A phase 2, randomised, double-blind, multi-site, placebo-controlled trial has been conducted to evaluate the efficacy, safety, and dose response of STS01 in mild to moderate patchy AA. The trial met its primary endpoint, with significantly more patients treated with 1% STS01 achieving a >30% Severity of Alopecia Tool (SALT) score improvement compared with placebo (75.9% vs 36.7% at 6 months; p=0.0037). Similar patterns of success were observed with >60% and 100% response criteria The trial included the assessment of patient-reported outcomes (PROs) using two validated AA-specific assessment scales: the Alopecia Areata Symptom Impact Scale (AASIS)^3,4^ and the Alopecia Areata Quality of Life Questionnaire (AAQLI). In this report, we present the analysis of the AASIS and compare findings with trials in severe AA including the pivotal phase 2b/3 trial for the JAK inhibitor ritlecitinib (ALLEGRO) which also used this scale.^4^

## Report

The study design has been previously published (NCT06402630/Eudract 2021-004145-20). Briefly, adult patients with mild to moderate AA (with guideline ≥10%–≤50% of scalp hair loss) for ≥6 months were eligible. Patients were randomly assigned to treatment with topical STS01 at doses of 0.25%, 0.5%, 1%, 2% or placebo, daily for 6 months. Overall, 155 patients received STS01 (n=30–31 across doses) or placebo (n=32), and baseline characteristics were similar between groups (Table S1– see supporting information).

The AASIS is a disease-specific tool that assesses the severity of AA symptoms and how symptoms interfere with daily functioning, consisting of 17 items across 3 subdomains, a global score and a total score (Figure 1). The scores were derived using an unweighted summing of the selected question scores. The AASIS was completed at baseline and at the 6-month visit (or earlier in the event of discontinuation), and derived total, global and subdomain scores were analysed using an ANCOVA model for change from baseline at endpoint as response variable, with treatment dose as fixed effect and baseline as covariate. As performed in the ALLEGRO trial, the correlation between AASIS and SALT scores at baseline and change from baseline scores was examined using Pearson’s correlation coefficients. In addition, an exploratory “shift” analysis was performed, whereby the ASSIS scores were divided into quartiles, and the proportion of patients in each quartile at baseline and end of treatment was determined, with “improvement” defined as at least a one quartile shift in the patient AASIS score at the end of treatment.

**Figure 1.**
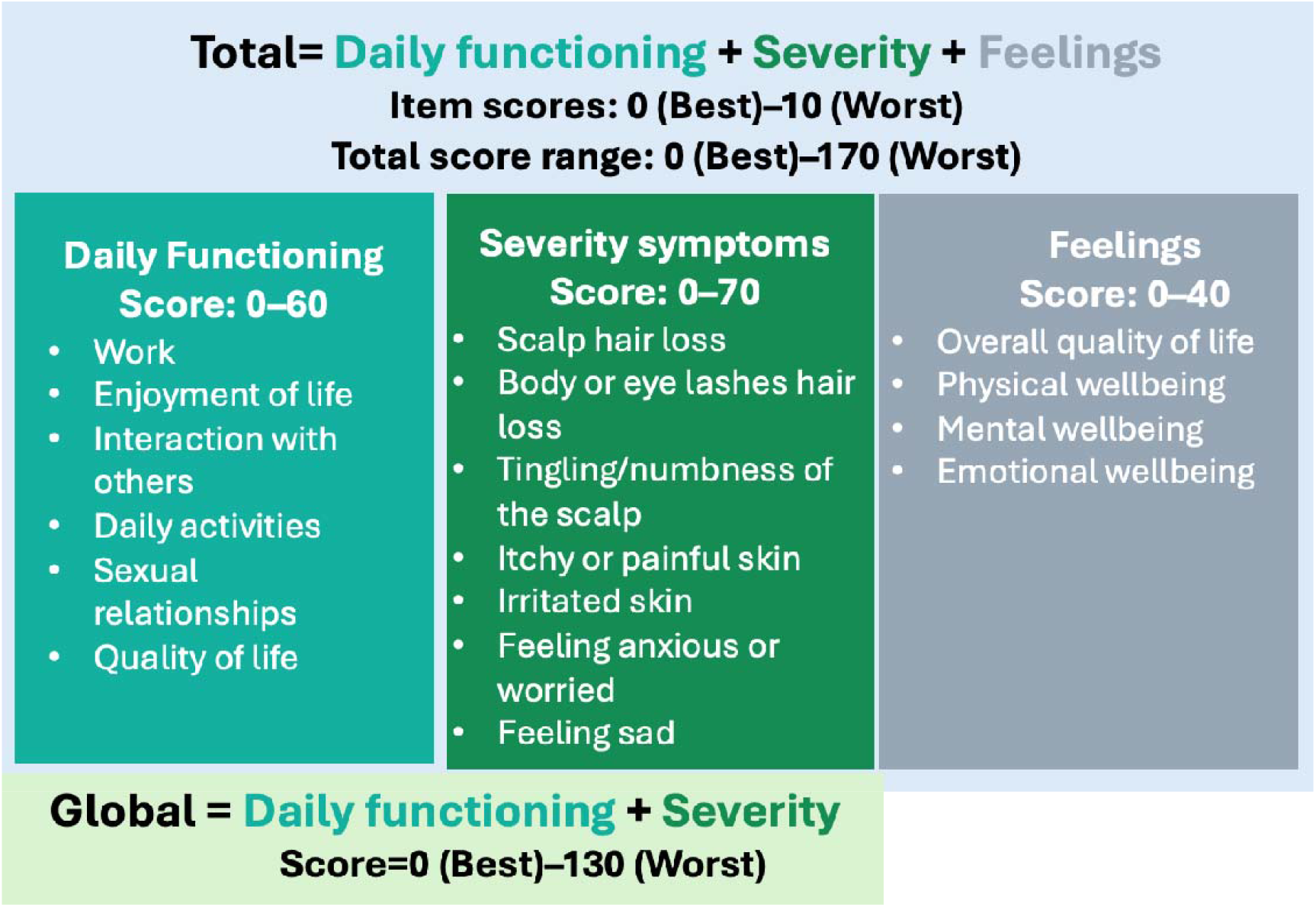
The AASIS assesses severity of AA symptoms and how symptoms interfere with daily functioning through a series of 17 questions each scored from 0 (not present) to 10 (as bad as you can imagine). The ASSIS consists of 3 domain scores (daily functioning [6 questions], symptom severity [7 questions], and feelings [4 questions]), a global score and a total score

Mean (standard deviation [SD]) AASIS total scores at baseline appeared reasonably comparable across the treatment groups (overall: 48.1 [31.3], ranging from STS01 2%: 30.7 [22.7] to STS01 0.5%: 58.6 [36.1]). With a possible score ranging from 0 (best) to 170 (worst) this suggests most patients were not particularly ‘troubled’ at the outset with a score less than one third of that possible (Table S1). There did however appear to be some correlation between AASIS total score at baseline and SALT scores (i.e severity of hair loss) at baseline although this was not pronounced (Pearson correlation 0.236) (Figure S1).

Although there was some evidence of a dose response increase in response from STS01 0.5% to 2%, the greatest improvements (decreases) in least squares (LS) mean changes from baseline were observed in the STS01 0.25% group for total score (-14.6), severity score (-4.3), global score (-12.7), and daily functioning score (-8.3) (Figure 2). From the hair regrowth efficacy perspective this was considered the minimally effective dose. However, some correlation was observed between AASIS change from baseline and SALT change from baseline, which was more pronounced with active treatment over placebo (Figure 3A). The shift analysis for the total score showed a trend towards the lower quartile categories at the end of treatment (Figure 3B; Table S2). More patients improved (defined as a reduction of 1 or more changes in quartiles) with all doses compared to placebo (42–50% with active treatment versus 22% with placebo) (Figure 3C). This improvement was similar across subgroups of patients including the mild and moderate subgroups, females, advancing AA condition and AA duration <4years, and therefore we could not identify predictors of HRQoL (Table S3).

**Figure 2.**
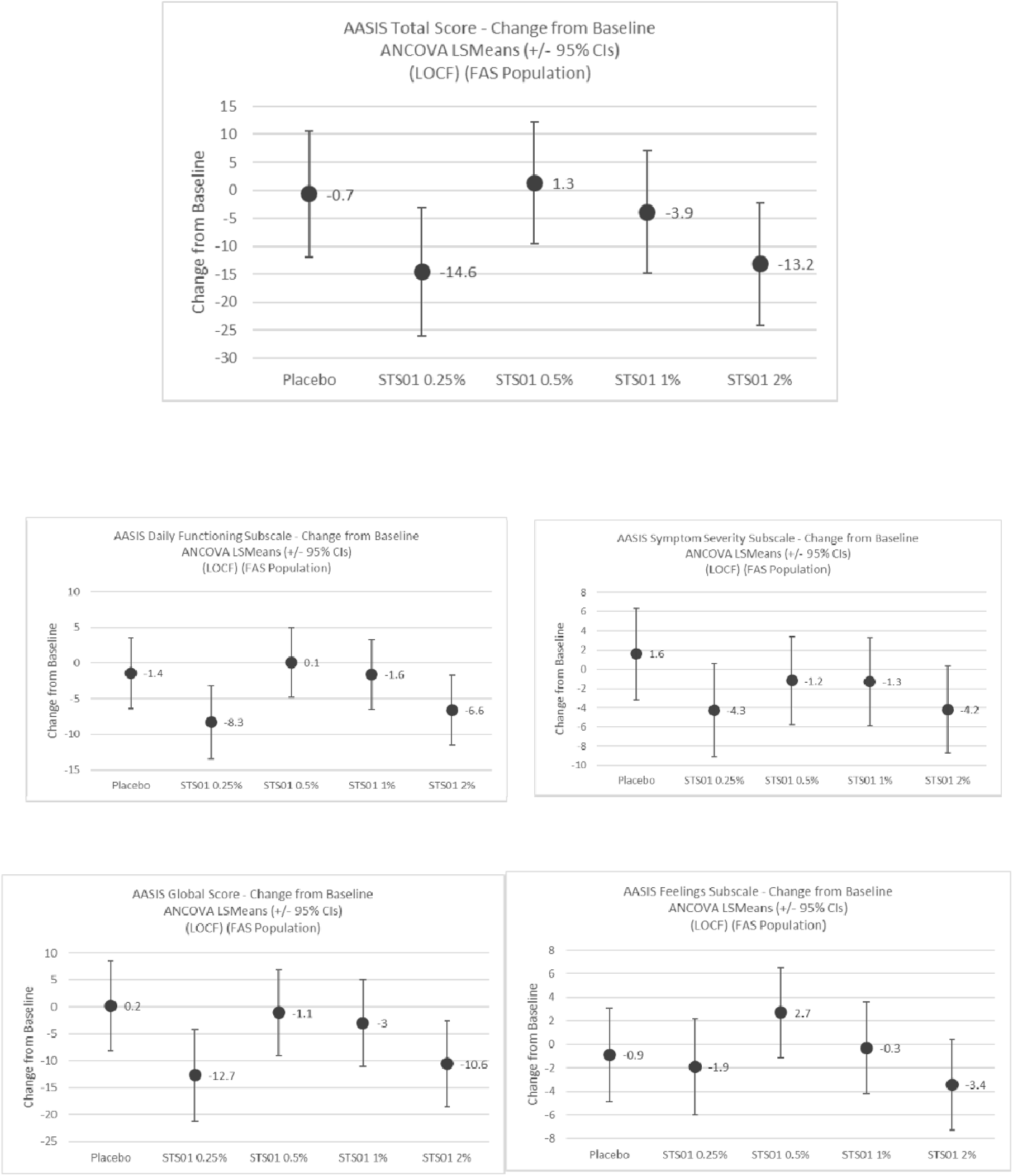
Change from baseline in AASIS scores for the total and global scores and the daily functioning, severity of symptoms and feelings domain scores

**Figure 3.**
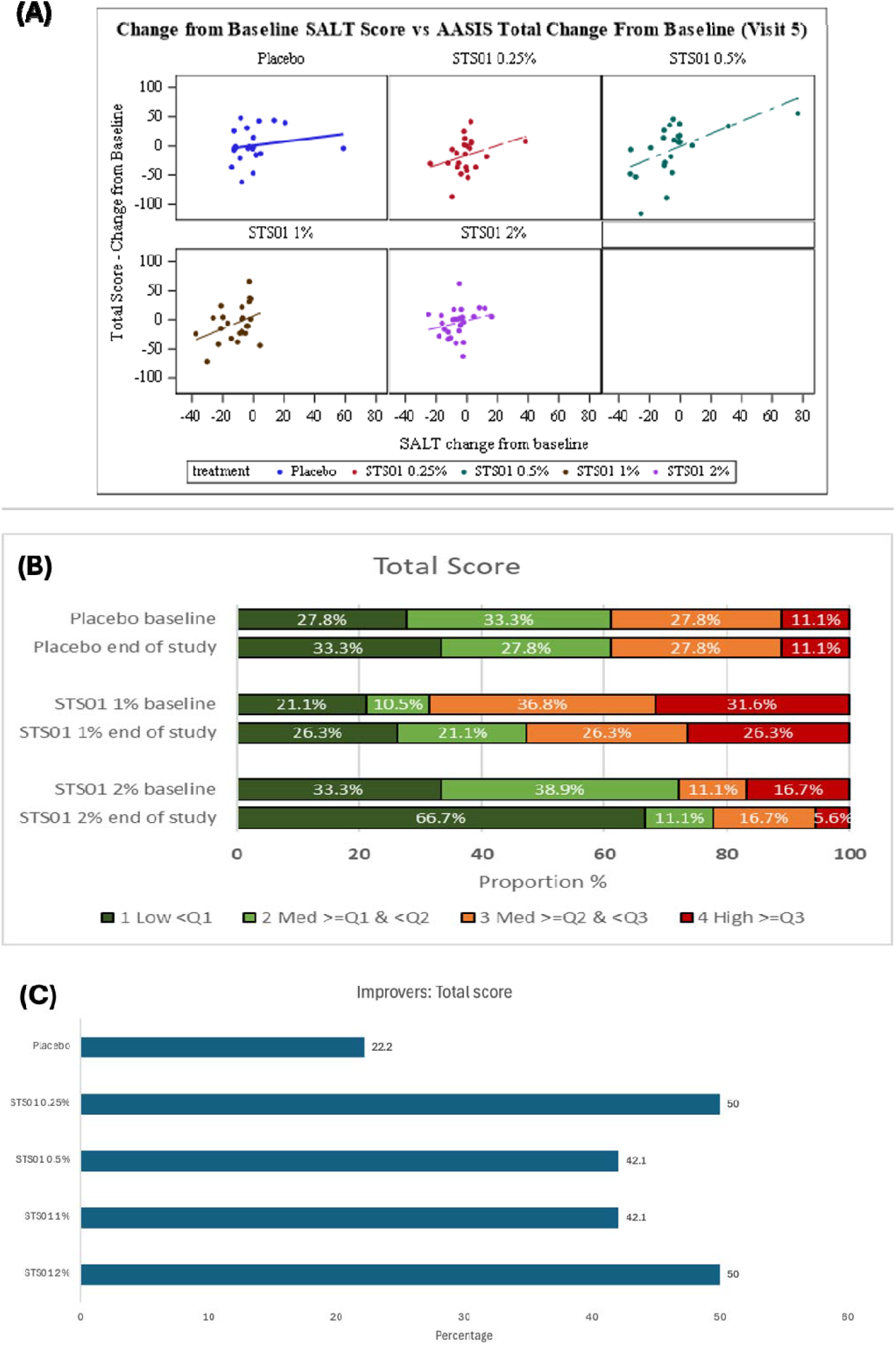
(A) Change from baseline is SALT scores vs. change from baseline in total AASIS score (B) Shift analysis of AASIS total score: proportion of patients in each AASIS quartile at baseline and end of treatment (C) AASIS total score: proportion of patients with improvement, defined as at least a one quartile shift in the patient AASIS score at the end of treatment

Overall, the baseline AASIS scores in this trial in mild to moderate AA were similar to baseline scores in severe AA trials (Table 1), suggesting comparable HRQoL burden irrespective of disease severity, with some correlation between changes in AASIS scores from baseline and clinical assessment SALT scores which appeared related to treatment. However, in both trials, significant improvements in hair regrowth did not appear to translate into significant HRQoL improvements (Table 1).

**Table 1.** AASIS comparison of baseline and change from baseline scores across clinical trials.

| AASIS Domain | Mean score at baseline in the total population |  |  |  |  |  |
| --- | --- | --- | --- | --- | --- | --- |
|  | SOT01<sup>a,b</sup><br>Mild to moderate<br>AA<br>N total=95 |  | ALLEGRO:<br>severe AA<br>N total=95 |  | Cyclosporin<sup>c</sup><br>Severe AA<br>N total=32 |  |
| Global Subscale (13 items) | 2.68 |  | 2.5 |  | Not done |  |
| Symptom Severity (7 items) | 2.63 |  | 3.0 |  | 3.9 |  |
| Daily Functioning (6 items) | 2.76 |  | 2.1 |  | 3.2 |  |
| Feelings (4 items) | 2.46 |  | Not done |  | Not done |  |
| AASIS Domain | Mean score change from baseline in the placebo and treatment groups |  |  |  |  |  |
|  | SOT01<sup>a,b</sup><br>Mild to moderate<br>AA |  | ALLEGRO<br>Severe AA |  | Cyclosporin<sup>c</sup><br>Severe AA |  |
|  | Placebo | STS01<br>1% | Placebo | Ritlecitinib | Placebo | Cyclosporin |
|  | N=18 | N=19 | N=47 | N=48 | N=16 | N=16 |
| Global Subscale (13 items) | 0.2 | -1.0 | 0.5 | -1.0 | Not done |  |
| Symptom Severity (7 items) | 0.2 | -0.6 | 0.1 | -1.5 | -0.4 | 0.1 |
| Daily Functioning (6 items) | -0.2 | -1.4 | 0.9 | -0.4 | -0.9 | -1.1 |
| Feelings (4 items) | -0.2 | -0.7 | Not done |  | Not done |  |

<sup>a</sup> Patients who completed 6 months of treatment
<sup>b</sup> Scores for the STS01 trial are approximated from: (mean total scores/total range score)\*10 to align with the scale of data from the ALLEGRO trial
<sup>c</sup> Scores for the cyclosporin trial are approximated from the 0-1 range reported by \*10 to align with the data from the ALLEGRO trial

As has been postulated from the other trials in severe AA, this may be related to limitations with the current measures used, which may not be capturing the HRQoL either at baseline, or in response to successful treatment. Certainly, in this respect, the baseline ASSIS scores were low in both our trial and in those in severe AA (Table 1). In addition, similar to the ALLEGRO trial, even the subgroup of patients classified with total hair regrowth did not demonstrate differences in AASIS scores compared to those without total regrowth. We did observe a ‘shift’ to the lowest quartile score with treatment – twice as many patients as placebo, however this did not translate to a particular patient type or demographic or regrowth subgroup. A recognised limitation of the AASIS is that patients did not have direct input during its development; however, even after development of a new patient-focused specific scale (Alopecia Areata Patient Priority Outcomes Instrument, AAPPO) evaluating hair loss, emotional symptoms and activity limitations,^5^ the ALLEGRO trial found no significant improvements in emotional symptoms or activity limitations from baseline or between groups (treatment vs. placebo).^6^

In developing new measures for use in future trials, there are several challenges that need to be acknowledged. Patients with AA may develop coping mechanisms to lessen the impact on lifestyle, especially if they have been dealing with AA for many years,^7^ as in this trial where the majority had had AA for ≥4 years. Furthermore, given that people with AA have been living with an unpredictable condition, the psychological effects, such as fear of recurrence and prior experiences of loss, can continue to impact feelings even after physical recovery, and patients may require time to ‘trust’ that their hair has regrown. As a consequence, even when hair regrows, improvements in HRQoL may take time to appear. Finally, AA is complex, with comorbidities including anxiety and depression that also impair HRQoL, which may be a result of the immune dysfunction as opposed to solely due to hair loss,^8^ and these co-morbidities may need to be addressed separately. A similar “psycholag’ (i.e a delay between physical and psychological improvement) has been observed in some people with psoriasis, with speculation that this phenomenon may involve immune dysregulation common to both the skin and psychiatric disease.^9^

Overall, the results from this study and others suggest the need for improved PRO tools to measure the impact of AA treatments. These could include the newly developed Scale of AA Distress (SAAD), a 41-item tool assessing the psychosocial impacts of AA,^10^ perhaps in conjunction with other tools such as the Hospital Anxiety Depression Scale (HADS). There should also be awareness of the potential for delays in reversing the psychosocial impacts, and assessments over a longer period may be necessary to reflect this. Finally, studies elucidating the intricate interplay between the psychological symptomatology, immune responses, and hair loss may provide well-needed insights in understanding the determinants of impaired HRQoL in individuals with AA.

## Data Availability

Provided on request

## Acknowledgements

Amanda Prowse PhD (Lochside Medical Communications) provided medical writing support.

## Data availability

The data underlying this article will be shared on reasonable request to the corresponding author.

## Ethics Statement

The study received approval from the responsible Independent Ethics Committee (London - South East Research Ethics Committee) (Reference: 21/LO/0851). All patients provided written informed consent.

## Supporting information

**Table S1.**
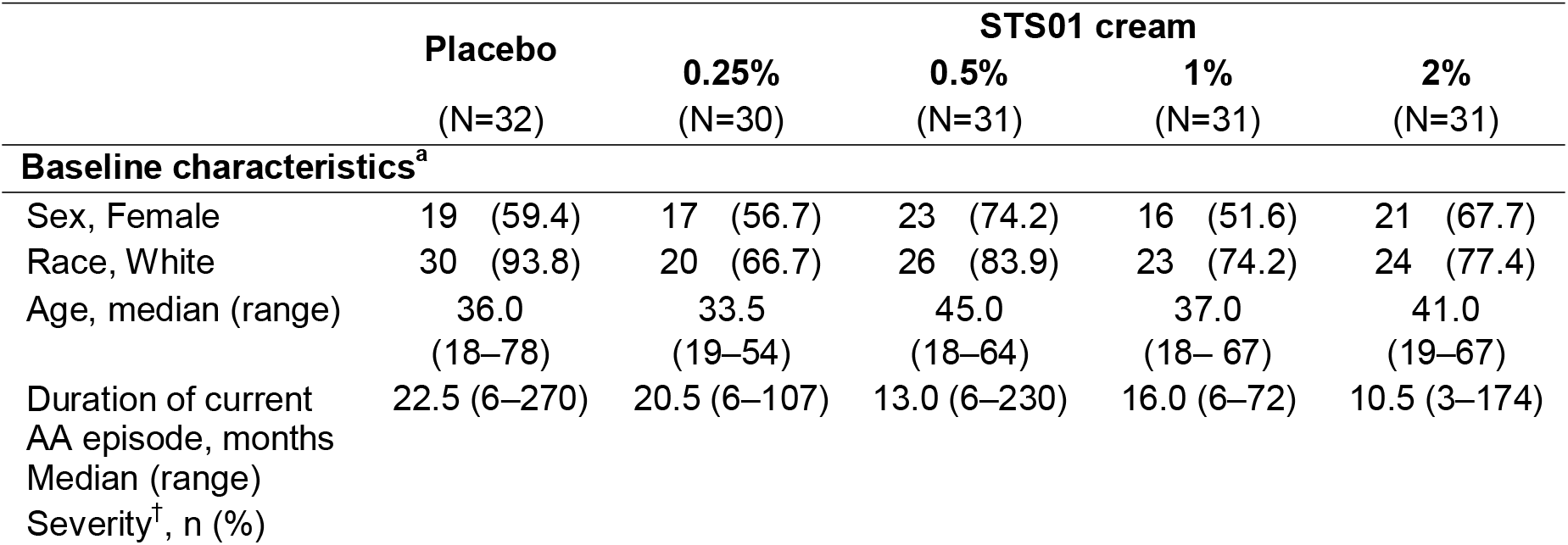
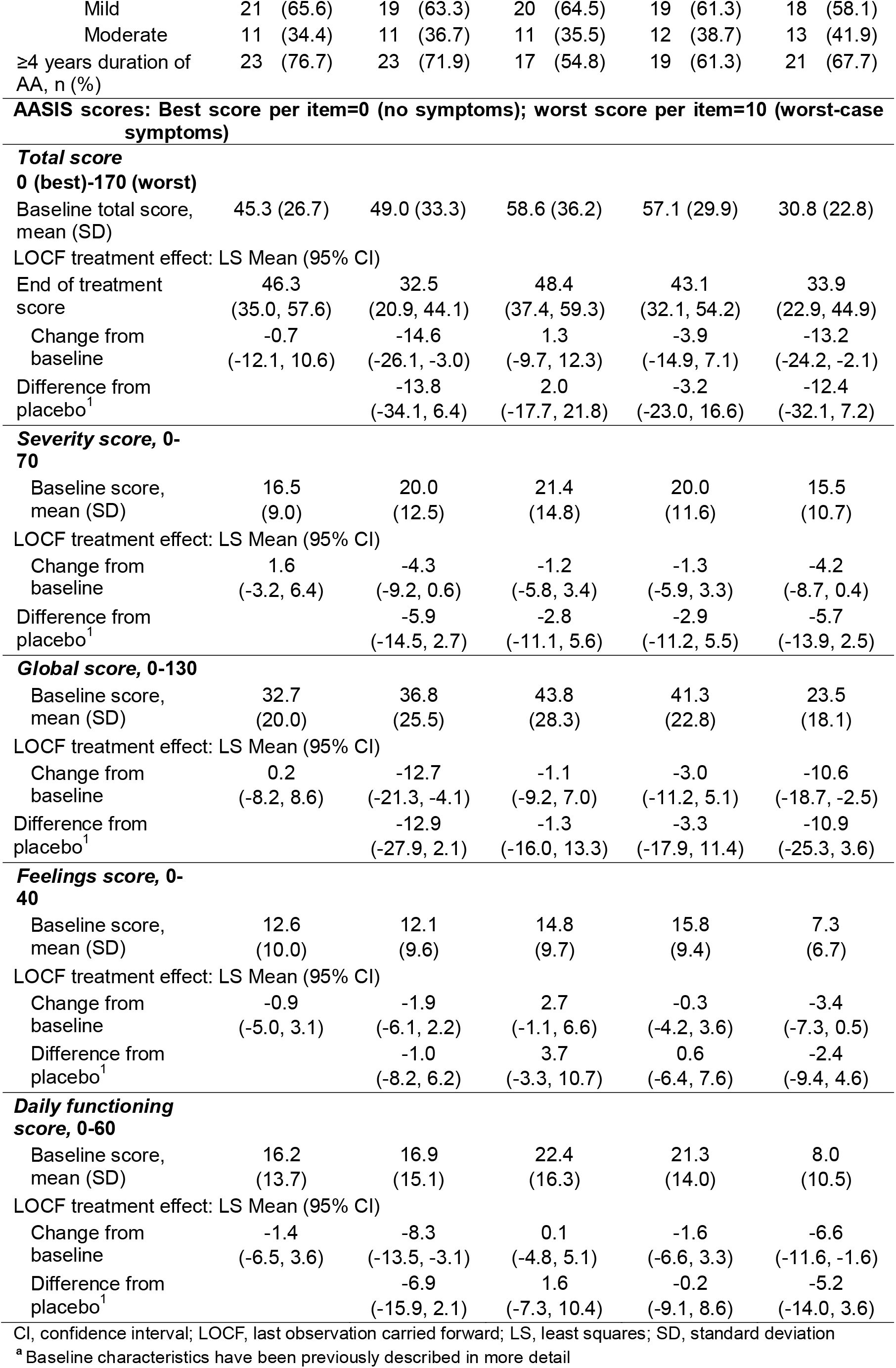
Baseline characteristics and AASIS scores (FAS population)

**Table S2.**
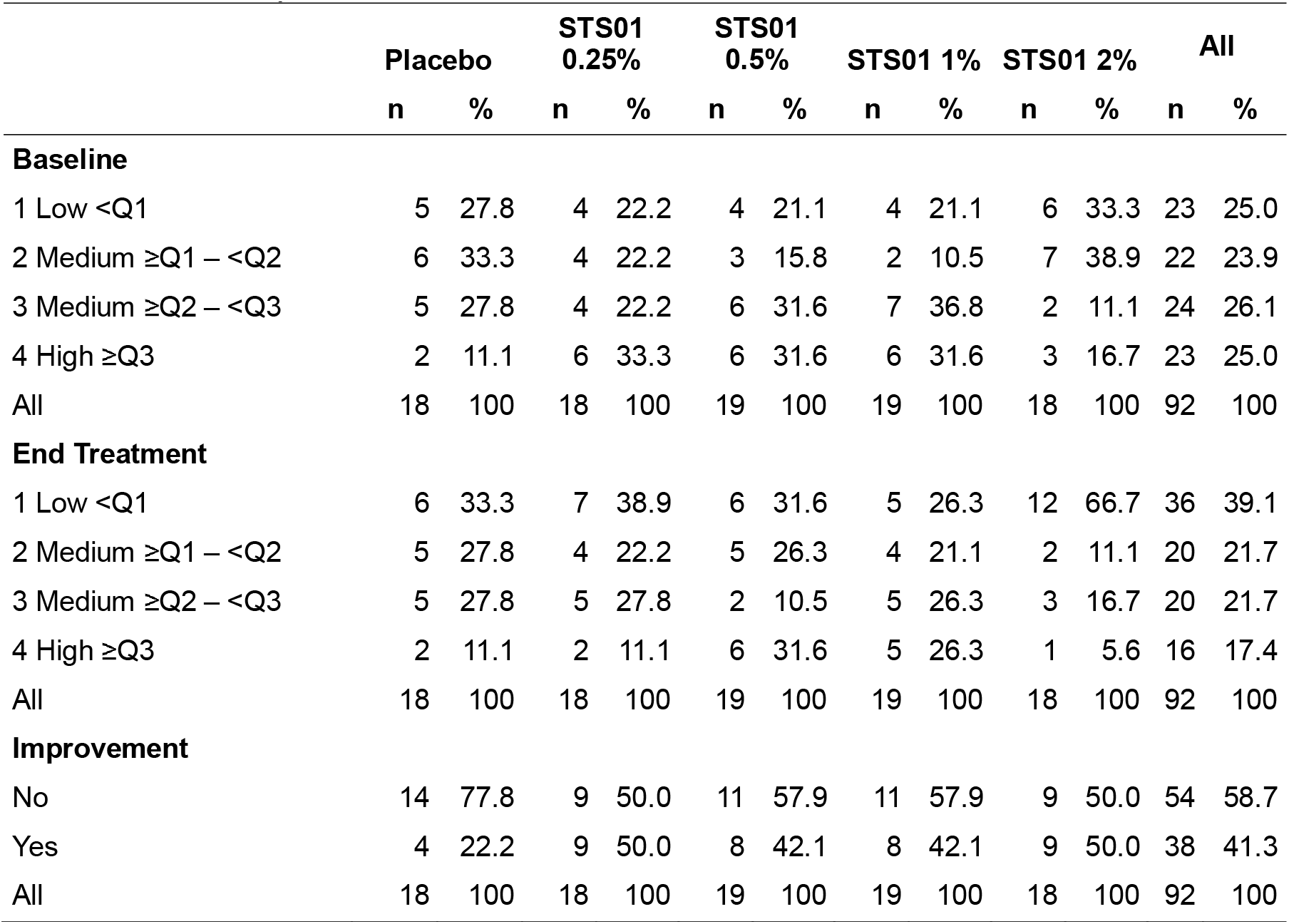
Shift analysis for AASIS total score.

|  | Placebo |  | STS01 0.25% |  | STS01 0.5% |  | STS01 1% |  | STS01 2% |  | All |  |
| --- | --- | --- | --- | --- | --- | --- | --- | --- | --- | --- | --- | --- |
|  | n | % | n | % | n | % | n | % | n | % | n | % |
| **Baseline** |  |  |  |  |  |  |  |  |  |  |  |  |
| 1 Low <Q1 | 5 | 27.8 | 4 | 22.2 | 4 | 21.1 | 4 | 21.1 | 6 | 33.3 | 23 | 25.0 |
| 2 Medium ≥Q1 – <Q2 | 6 | 33.3 | 4 | 22.2 | 3 | 15.8 | 2 | 10.5 | 7 | 38.9 | 22 | 23.9 |
| 3 Medium ≥Q2 – <Q3 | 5 | 27.8 | 4 | 22.2 | 6 | 31.6 | 7 | 36.8 | 2 | 11.1 | 24 | 26.1 |
| 4 High ≥Q3 | 2 | 11.1 | 6 | 33.3 | 6 | 31.6 | 6 | 31.6 | 3 | 16.7 | 23 | 25.0 |
| All | 18 | 100 | 18 | 100 | 19 | 100 | 19 | 100 | 18 | 100 | 92 | 100 |
| **End Treatment** |  |  |  |  |  |  |  |  |  |  |  |  |
| 1 Low <Q1 | 6 | 33.3 | 7 | 38.9 | 6 | 31.6 | 5 | 26.3 | 12 | 66.7 | 36 | 39.1 |
| 2 Medium ≥Q1 – <Q2 | 5 | 27.8 | 4 | 22.2 | 5 | 26.3 | 4 | 21.1 | 2 | 11.1 | 20 | 21.7 |
| 3 Medium ≥Q2 – <Q3 | 5 | 27.8 | 5 | 27.8 | 2 | 10.5 | 5 | 26.3 | 3 | 16.7 | 20 | 21.7 |
| 4 High ≥Q3 | 2 | 11.1 | 2 | 11.1 | 6 | 31.6 | 5 | 26.3 | 1 | 5.6 | 16 | 17.4 |
| All | 18 | 100 | 18 | 100 | 19 | 100 | 19 | 100 | 18 | 100 | 92 | 100 |
| **Improvement** |  |  |  |  |  |  |  |  |  |  |  |  |
| No | 14 | 77.8 | 9 | 50.0 | 11 | 57.9 | 11 | 57.9 | 9 | 50.0 | 54 | 58.7 |
| Yes | 4 | 22.2 | 9 | 50.0 | 8 | 42.1 | 8 | 42.1 | 9 | 50.0 | 38 | 41.3 |
| All | 18 | 100 | 18 | 100 | 19 | 100 | 19 | 100 | 18 | 100 | 92 | 100 |

**Table S3.**
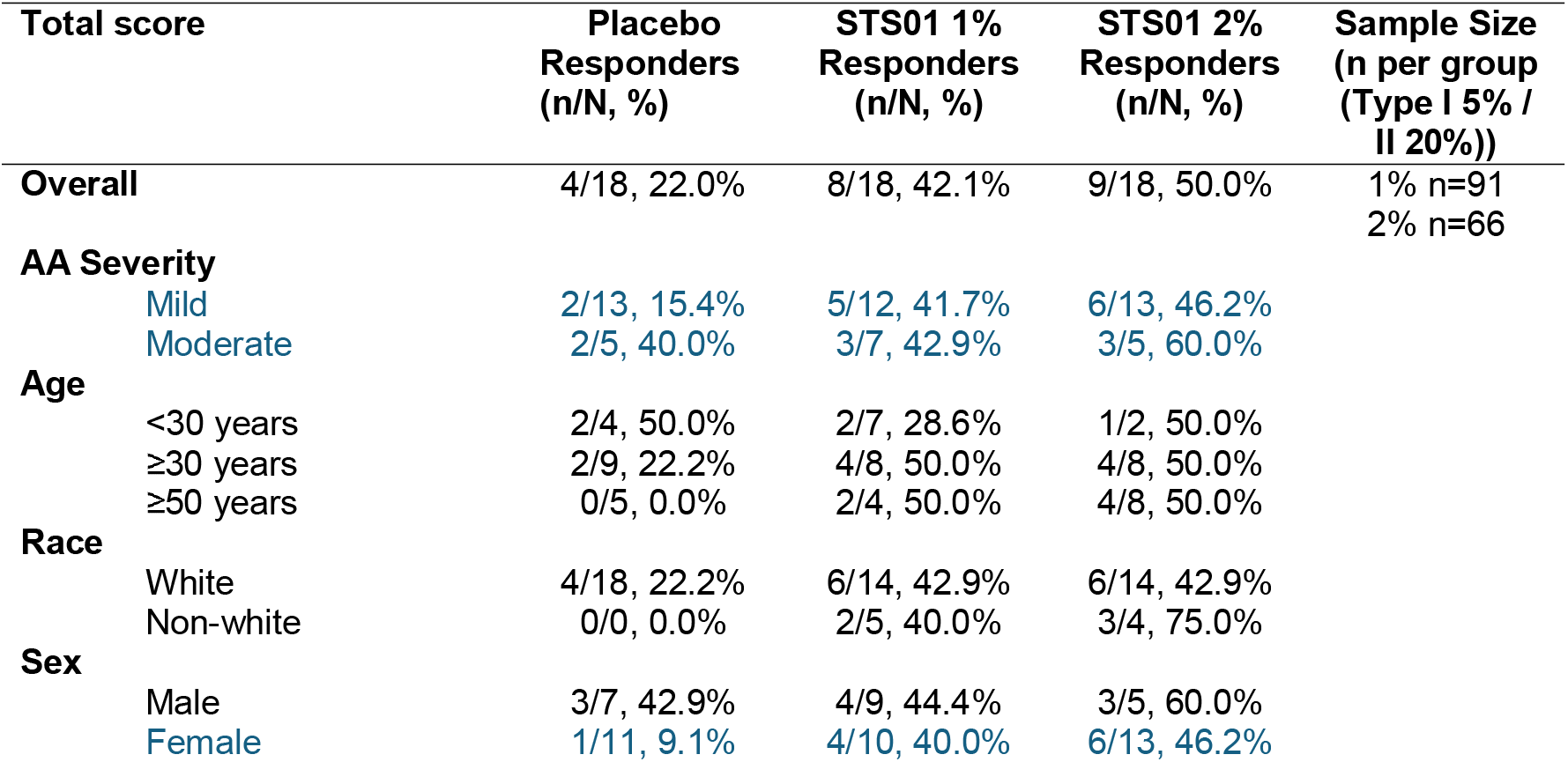
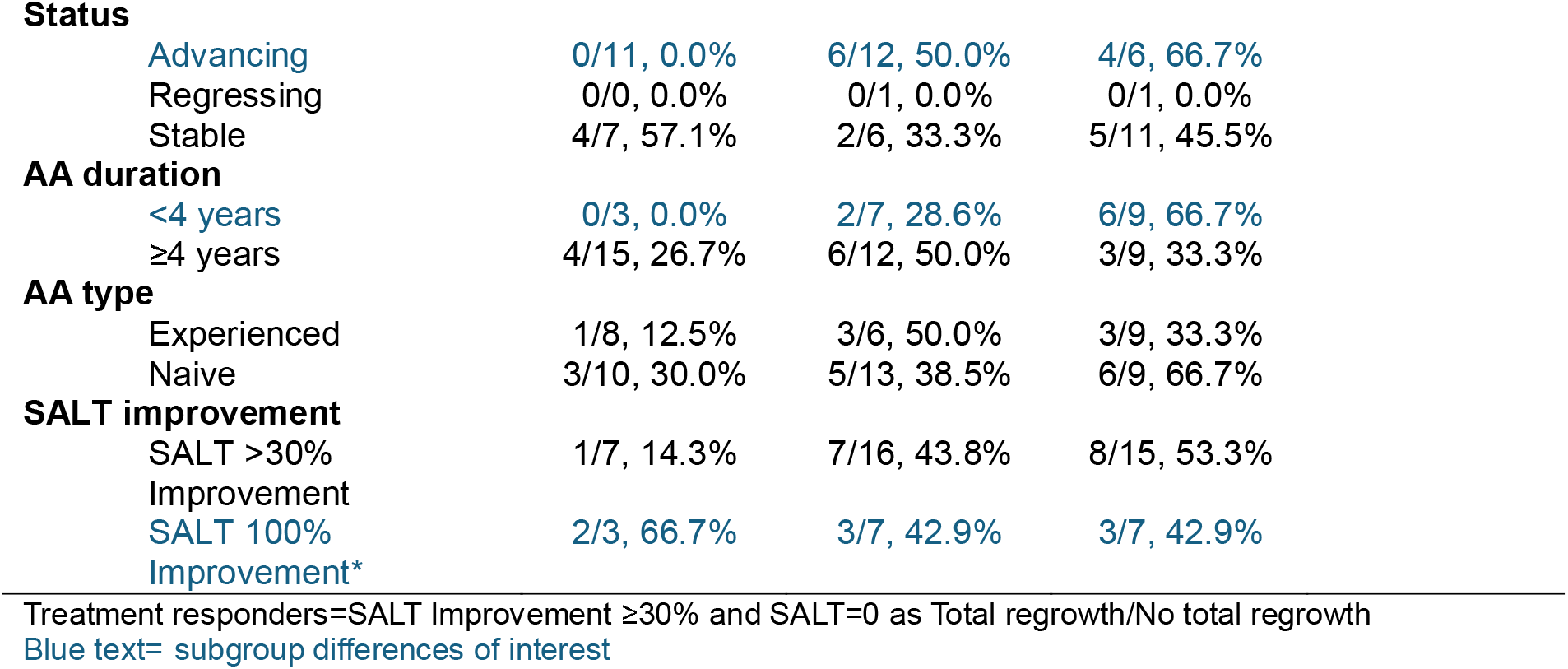
Overall summary of responders as defined by AASIS QoL quartile decrease after treatment.

**Figure S1.**
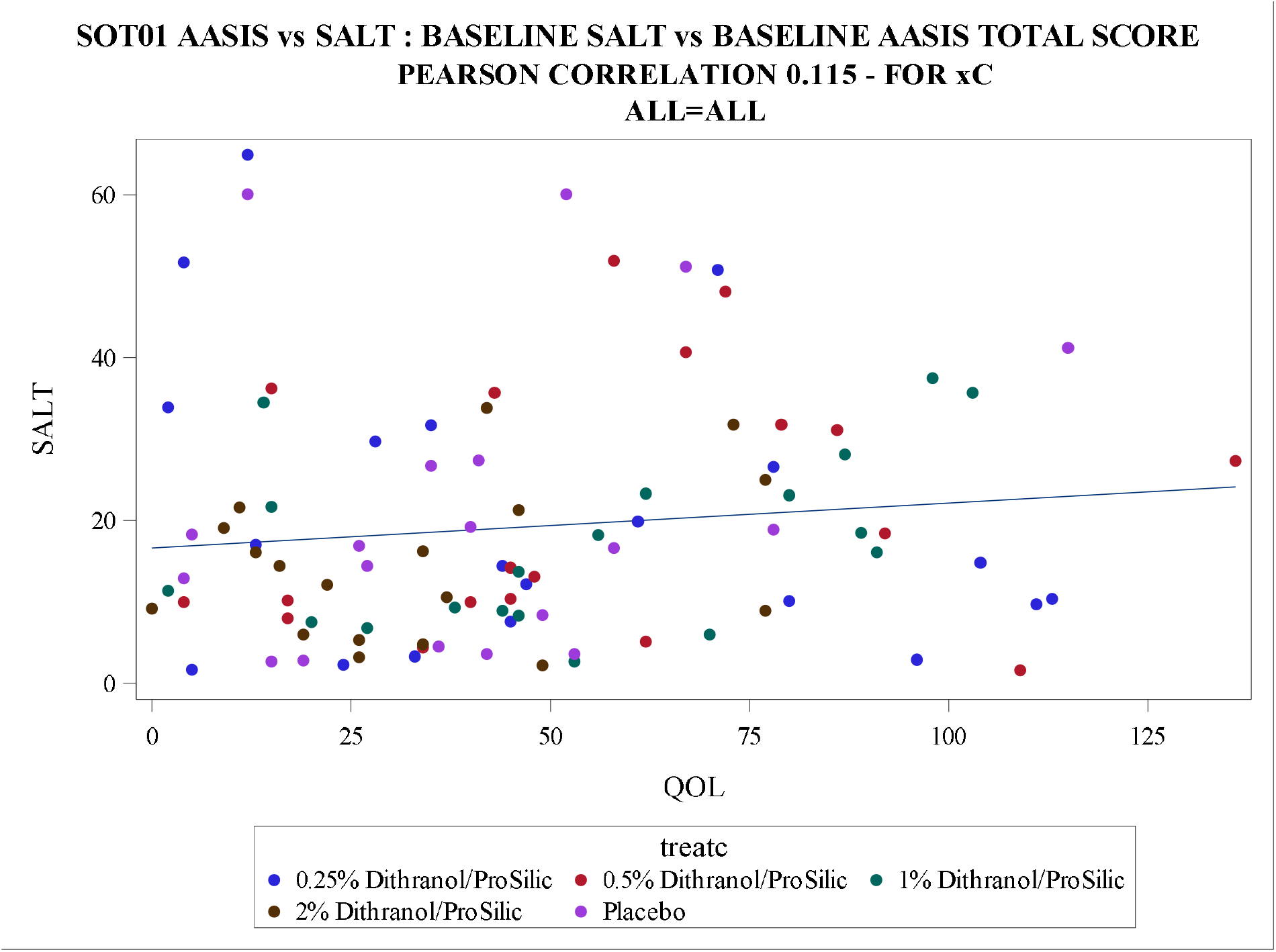
Baseline SALT scores vs. baseline total AASIS score

## References

1 Aldhouse NVJ, Kitchen H, Knight S, Macey J, Nunes FP, Dutronc Y, Mesinkovska N, Ko JM, King BA, Wyrwich KW. “‘You lose your hair, what’s the big deal?’ I was so embarrassed, I was so self-conscious, I was so depressed:” a qualitative interview study to understand the psychosocial burden of alopecia areata. Journal of Patient-Reported Outcomes 2020; 4: 76.

2 Gelhorn HL, Cutts K, Edson-Heredia E, Wright P, Delozier A, Shapiro J, Senna M, Tosti A. The Relationship Between Patient-Reported Severity of Hair Loss and Health-Related Quality of Life and Treatment Patterns Among Patients with Alopecia Areata. Dermatol Ther (Heidelb) 2022; 12: 989–97.

3 Lai VWY, Chen G, Sinclair R. Impact of cyclosporin treatment on health-related quality of life of patients with alopecia areata. J Dermatolog Treat 2021; 32: 250–7.

4 Winnette R, Banerjee A, Sikirica V, Peeva E, Wyrwich K. Characterizing the relationships between patient-reported outcomes and clinician assessments of alopecia areata in a phase 2a randomized trial of ritlecitinib and brepocitinib. J Eur Acad Dermatol Venereol 2022; 36: 602–9.

5 Winnette R, Martin S, Harris N, Deal LS. Development of the Alopecia Areata Patient Priority Outcomes Instrument: A Qualitative Study. Dermatol Ther (Heidelb) 2021; 11: 599–613.

6 Sinclair R, Mesinkovska N, Mitra D, Wajsbrot D, Law EH, Wolk R, King B. Patient-Reported Hair Loss and Its Impacts as Measured by the Alopecia Areata Patient Priority Outcomes Instrument in Patients Treated with Ritlecitinib: The ALLEGRO Phase 2b/3 Randomized Clinical Trial. Am J Clin Dermatol 2025; 26: 109–19.

7 Welsh N, Guy A. The lived experience of alopecia areata: a qualitative study. Body Image 2009; 6: 194–200.

8 Bain KA, McDonald E, Moffat F, Tutino M, Castelino M, Barton A, Cavanagh J, Ijaz UZ, Siebert S, McInnes IB, Astrand A, Holmes S, Milling SWF. Alopecia areata is characterized by dysregulation in systemic type 17 and type 2 cytokines, which may contribute to disease-associated psychological morbidity. Br J Dermatol 2020; 182: 130–7.

9 Ahmed A, Mohandas P, Taylor R, Assalman I, Bewley A. ‘Psycholag’: a new term to describe the delay between physical and psychological improvement in patients with skin disease. Br J Dermatol 2024; 192: 1–3.

10 Gorbatenko-Roth K, Wood S, Johnson M, Wallander I, Nugent J, Hordinsky M. Beyond health-related quality of life: initial psychometric validation of a new scale for addressing the gap in assessing the full range of alopecia areata psychosocial burden. Br J Dermatol 2023; 189: 71–9.

